# Analysis of the association between hand grip strength and functionality in community elderly

**DOI:** 10.1101/2020.07.14.20153411

**Authors:** Mariana Edinger Wieczorek, Cislaine Machado de Souza, Patrícia da Silva Klahr, Luis Henrique Telles da Rosa

## Abstract

**Objective:** To analyze an association between handgrip strength (HGS) and functional performance tests by healthy, non-institutionalized elderly.

**Method:** This is a cross-sectional study. A sample consisted of 36 elderly people (66.69 ± 4.84 years), all of whom responded to a cognitive assessment instrument and another to assess the level of physical activity, in addition to sociodemographic data and health conditions. The HGS was measured using the hydraulic dynamometer JAMAR and performed the six-minute walk test (6MWT) and the Timed Up and Go test (TUG) to assess the cardiorespiratory capacity submitted during displacement and body mobility.

**Results:** It was possible to verify through the Pearson coefficient the weak and significant association between the MPH and the variables 6MWT (p≤0.05) and TUG (p = 0.027).

**Conclusion:** For samples of healthy elderly and applied experimental conditions, the FPM is related to performance tests. Thus, it is believed that the evaluation of the HGS may be an alternative to interference in this population.

## INTRODUCTION

The aging process is naturally accompanied by several multifactorial and progressive changes. Among these, not only physiological and metabolic transformations stand out, but also structural and functional. The declines resulting from the aging process, as well as the dysfunctions and diseases prevalent in this age group, may compromise the functionality, independence and quality of life of the elderly.^1-4^

The loss of muscle mass and consequently muscle strength and power is the main responsible for the deterioration in mobility and functionality of the aging individual.^2-4^ The elderly present a decrease in muscle strength from muscular, neurological, endocrine or environmental mechanisms.^2,5^ The decline in strength and mobility of the elderly interferes with the performance of activities essential for functional performance., such as walking, sitting, standing, climbing stairs, among others.^1,4^

As a general indicator of muscle strength and power, in clinical practice, handgrip strength (HGS) has been used.^6^ In addition, the evaluation of HGS is used as an indicator of global strength^8,9^ and functionality^7,8^, however, not all studies have been able to demonstrate the existence of significant correlations between HGS and functional performance.^10^ There are several ways to assess the functional status and level of physical fitness of the elderly., often by inference, because it is known that the worse the health status, the greater the negative impact on mobility, the ability to perform a submemal task and the independence of the elderly.^13,14^

Usually for the execution of functional tests, muscle groups essential for body weight support are recruited, such as trunk and lower limb stabilizing muscles^10,11,12^, the performance is due to the ability to generate strength in a given test time (slow contraction fibers, type I), while the HGS assesses the peak muscle strength of upper groups (fast contraction fibers - type II), more resistant to fatigue.^11,12^

In general, functional performance tests involve tasks that require postural transfer, mobility and displacement.^12,13^ The six-minute walk test (6MWT) measures the maximum distance traveled in the period of 6 minutes, being commonly used to assess submemal cardiorespiratory capacity during activities that require displacement.^13,14^ The Timed Up and Go (TUG) test indicates functional mobility, whose performance is related to gait, postural and steering changes during the act of walking, being evaluated through the time spent performing the course.^12,13^

There are still gaps regarding the relationships between HGS and the functionality of the elderly.^10^ The reproducibility of HGS as an indicator of functional performance in people over 60 years of age is questionable. This study aimed to verify the association between HGS, submemal cardiorespiratory capacity during commuting and body mobility in healthy, non-institutionalized elderly.

## METHODOLOGY

This is an observational study with a cross-sectional design, approved by the Research Ethics Committee of the Federal University of Health Sciences of Porto Alegre by protocol number 2.137.840/2017. This study integrates a larger research on the theme of functionality in the elderly, developed in the institution by the Grupo de estudos em Reabilitação (GEReab).

The subjects were deceived by means of social media and local dissemination actions with flyers and posters. We included elderly aged 60 years or older living in the city of Porto Alegre, RS, Brazil. For inclusion criteria, healthy elderly were excluded, those who presented autonomy and independence and control of chronic diseases and physical symptoms.^15,16^ The elderly were excluded in the postoperative period of any nature, with diseases or physical deficits and who characterize a pathological aging process and that prevents the performance of the tests.

The sociodemographic and clinical variables of interest were: gender, age, weight and height, body mass index, schooling, health problems, medication use, occupational, social activities and physical exercise. The Mini Mental State Examination (MMSE)^17^ was applied for the cognitive knowledge of the elderly, using the following cut-off points: 18 points for illiterate, 21 for those who were educated with one and three years, 24 for individuals between four and seven years of formal education, and 26 for people with more than seven years of schooling. The International Physical Activity Questionnaire (IPAQ)^18^, in a short form, was applied to assess the level of physical activity. This version presents seven questions, whose information estimates the time spent per week in different areas of physical activity, such as work, means of transportation, domestic activities; of recreation, sport or leisure, and sitting time spent.^19^ The final classification determines how sedentary, irregularly active, active and very active.^20^

### Data collection

To ensure the quality of the evaluation protocol, the researchers responsible for data collection were properly trained. Both the initial interview and the protocol for the application of the tests to the individuals included in the research took place at the Physiotherapy Laboratory of the Federal University of Health Sciences of Porto Alegre, from January 2018 to July 2019. The evaluation of handgrip strength and functional tests were applied on the same day.

### Handgrip Force (HGS)

To measure handgrip strength, a portable hydraulic dynamometer of the JAMAR brand was used.^21^ The measurement was verified with the individual sitting in a chair with backrest and without armrests. The shoulder of the tested limb was adduced and in neutral rotation, elbow in flexion of 90 degrees, forearm in neutral position and wrist between 0 and 30 degrees of extension and between 0 and 15 degrees of ulnar deviation.^6,21,22^ A demonstration of how the test should be performed for familiarization with the equipment and participants made a simulation was performed. During the test, there was encouragement through standardized verbal command. Participants were instructed to perform maximum contraction using the dominant hand.^6,22^ Three measurements were collected, assuming the best performance among the three attempts as a reference^6,22^, there was a 30-second rest interval between each test.^21,22^ Some factors may hinder the establishment of normative values of HGS, among them can be mentioned: gender, age, dominance, evaluation time, body positioning and anthropometric characteristics.^6,21,22^

### Six-Minute Walk Test (6MWT)

The TC6 was applied as recommended by the American Thoracic Society^23^, briefly the subjects were instructed to walk from side to side in a corridor of 30 meters, for a period of 6 minutes, being instructed to walk as fast as possible, without running. Before the beginning of the test, blood pressure (BP), peripheral oxygen saturation (SpO2), Borg scale, heart rate (HR) and respiratory rate (RR) were measured. The variables were collected again after the end of the test. The evaluator walked throughout the test a little behind the individual, who was monitored throughout the period by the oximeter. During the test, all participants were verbally encouraged in a pre-established manner, every minute elapsed. Those who felt tired could slow down the walk or even stop if necessary, and the timer was not stopped during rest. After 6 min, the subject was instructed to stop and the distance covered on the last lap was measured.

### Timed Up and Go Test (TUG)

A chair with backrest was positioned and the elderly man was asked to get up from the chair, without hand support, to travel a distance of 3 meters, to turn around and to return by sitting in the chair again. During the journey the stopwatch was started when the elderly man got up from his chair, and interrupted when his back resumed support on the back of the chair. TUG time was measured in seconds (s). A practical essay was conducted with each participant to become familiar with the task.^24^

### Statistical analysis

Parametric numerical data were expressed as mean and standard deviation. Initially, the Shapiro-Wilk test was performed to verify the normality of the data. The descriptions of the qualitative variables were expressed in absolute and relative frequency. Pearson’s correlation coefficient was used to verify the correlation between HGS and the 6MWT and TUG functional performance tests. To compare the other variables between the tests, the Student’s t-test was used for dependent samples. The level of significance adopted was (p≤ 0.05).

## RESULTS

**Table 1** presents the characteristics of the sample studied. All participants in the survey reported being retired or pensioners, of which 58.33% reported having another occupation and 8.33% had an informal job. The 72.22% of elderly people who participated in social activities reported mostly religious commitments and participation in craft groups. According to the IPAQ questionnaire classification, the elderly identified as irregularly active A should reach at least the frequency of 5 days per week or the duration of 150 minutes per week, while the irregularly active B did not meet any of the criteria regarding frequency and duration.^20^

**Table 1.**
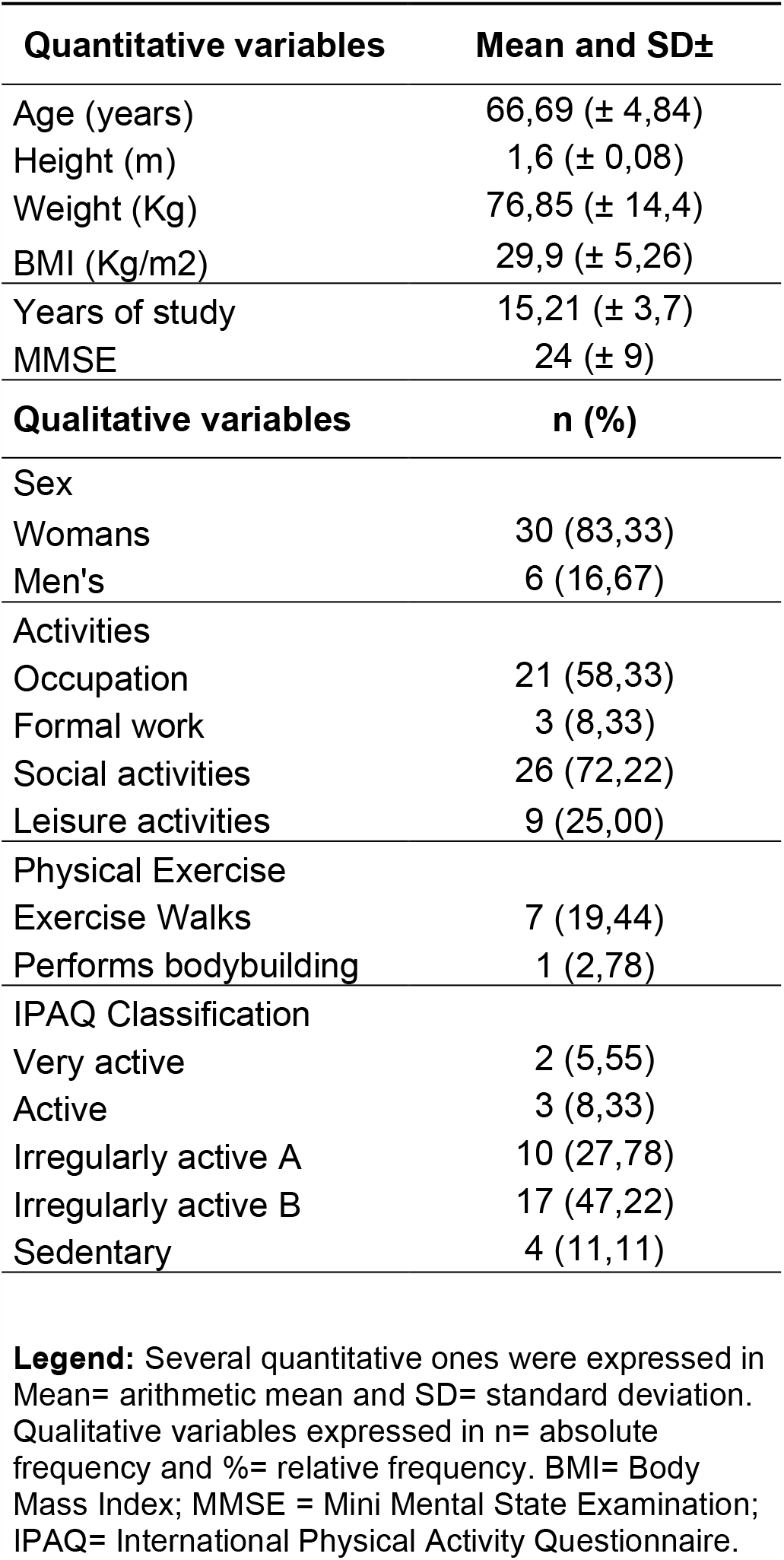
Characterization of the sample of healthy elderly (N=36). Porto Alegre, RS, 2019.

The results referring to the applied tests are presented in **Table 2**. All elderly participants were able to complete the functional performance tests without interruptions. **Table 3** shows the variables analyzed in the 6MWT, before and after the test, the values show that there was adequate response to exercise.

**Table 2.** Handgrip Strength results and functional performance tests. Porto Alegre, RS, 2019.

| Testes | Média e DP± |
| --- | --- |
| HGS (kgf) | 30,22 (± 8,36) |
| 6MWT (m) | 455,17 (± 90,41) |
| TUG (s) | 6,89 (± 1,6) |

**Legenda:** HGS= handgrip force; 6MWT= 6 minute walk test; TUG= *Timed Up and Go*.

**Table 3.** Variables of the 6-minute walk test in the pre- and post-test moments. Porto Alegre, RS, 2019.

| Variable | Pre-test variables<br>Mean and SD± | Post-test<br>Mean and SD± | p |
| --- | --- | --- | --- |
| SpO2 (%) | 97 (± 1,84) | 97,68 (± 1,20) | 0,010 |
| HR (bpm) | 77,15 (± 10,68) | 82,5 (± 10,62) | 0,004 |
| SBP (mmHg) | 131,88 (± 16,71) | 140,35 (± 19,79) | 0,003 |
| DBP (mmHg) | 77,5 (± 9,77) | 82,18 (± 10,01) | 0,003 |
| RR (irpm) | 17,5 (± 2,69) | 20,56 (± 3,26) | 0,000 |

**Legend:** SpO2= Oxygen saturation; HR= heart rate; SBP= systolic blood pressure; DBP= diastolic blood pressure; RR= respiratory rate; p= significance level.

**Table 4** shows the correlation between the HGS and the results of the 6MWT (distance traveled in meters) and in the TUG (time of displacement in seconds). The results using pearson’s coefficient revealed the existence of a weak but significant correlation between HGS and functionality, measured through functional performance tests.

**Table 4.** Correlation between handgrip strength and functional performance tests. Porto Alegre, RS, 2019.

| Correlation Test de Pearson | Handgrip Strength |  |
| --- | --- | --- |
|  | r | p |
| 6MWT | 0,324 | 0,05 |
| TUG | -0,385 | 0,027 |

**Legenda:** 6MWT= 6 minute walk test; TUG= *Timed Up and Go*; r = Pearson correlation; p = significance level.

## DISCUSSION

The aim of this study was to analyze the correlation between HGS and functional performance tests by healthy elderly, pearson’s correlation coefficient was used. The study demonstrated the weak and statistically significant association between the HGS and the variables related to the functional performance tests, and it was verified that the greater the number of meters traveled in the 6MWT and the shorter the time of performance in the TUG, the higher the HGS.

Regarding the characteristics of the sample, there was a predominance of elderly females 30 (83.33%), confirming the Brazilian demographic pattern, in which the absolute number of elderly women is higher than men.^25^ Only 3 (8.33%) of the elderly were classified as physically active and 2 (5.55%) as many assets; and the mean Body Mass Index (BMI) was 29.9 Kg /m^2^ (± 5.26). According to Lenardt et al.^26^, elderly men and women with lower levels of physical activity also have lower muscle mass and consequently a higher prevalence of physical disability.

Collaborating in this logic, Martin et al.^27^ discuss that physical inactivity can contribute to the functional loss of the elderly, due to physiological alteration of muscle mass and strength, decreasing fitness and physical performance. In this study, elderly people with BMI between 28 and 30kg/m^2^ were classified as overweight. The study by Furtado et al.^28^, analyzing BMI and HGS values, revealed that in this age group, the increase in BMI may be related to the decrease in strength in both genders. However, in this study with healthy elderly, compared with cutoff points already established in the literature, the mean values of HGS and performance tests were considered relatively high for this population.

This is probably due to the fact that most studies that evaluated these variables, in general, include frail elderly, comorbidity patients or institutionalized. A significant number of elderly in the study, 26 (72.22%), participated in social activities and 21 (58.33%) performed some occupation, which can indirectly predict a greater physical capacity, thus increasing the HGS and performance in the tests. The performance of physical activity in a systematic way contributes positively to higher levels of HGS^27,29^ and physical fitness^26^, however an active behavior in other domains and activities, even if not structured, but performed spontaneously throughout the day, especially in leisure^29^, can positively influence the same way^30^.

In the present study, the mean HGS was 30.22 Kgf (± 8.36). Based on the latest review of the European consensus on the definition and diagnosis of sarcopenia, the European Working Group on Sarcopenia in Older Persons (EWGSOP)31 redefined, through HGS results, the cutoff points related to sarcopenia (<27kg for men and <16kg for women), based on this study, the elderly in the sample did not present sarcopenia. According to Lenardt et al.^32^ cutoff points are important to predict disability, morbidity and mortality, mainly associated with frailty in the elderly.

Considering the results in relation to the values of the functional performance tests, for the 6MWT the distance measurement of 455.17m (± 90.41) was found. According to Britto and Sousa^33^, there are, in the literature, some formulas that can predict, based on sex, weight, height and age, the walking distance expected during the 6MWT, but it is assumed that healthy people can walk during the test, distances ranging between 400 and 700 meters; in unhealthy individuals, the distance walked during the test, less than 300 meters may be correlated with a high probability of death and/or hospitalization.

Regarding TUG, the time measurement found in the study was 6.89s (± 1.6). In the study by Rodrigues et al.^34^, a time of up to 10 seconds was pointed out, being considered normal, from 10.01 to 20 seconds representing impaired performance and above 20.01 seconds a higher risk of falls in the elderly, another measure of TUG was also suggested, taking into account age stratification. According to Zazá et al.^35^, it was verified that the performance of elderly people considered healthy and physically active does not differ from the reference values suggested for the TUG test.

Analyzing the results of the present study, we can verify that the measure of HGS can be associated with the functional performance of healthy and non-institutionalized elderly. Other studies have already demonstrated positive associations between HGS and functional tasks. In the study by Geraldes et al.^10^, a moderate correlation was found between HGS and performance in motor tasks. This association was stronger with all tests than with each of them taken alone. This result is due to the fact that the study probably also contains function tests with fine motor skills, such as taking and placing the key in a lock and taking out and replacing a lamp in a nozzle, for example.

Similarly, in the study by Oliveira et al.^7^, it was found that the HGS serves as an indicator of functionality, related to ADL,s and IADL,s. The results were through the application of the Barthel Index and the Lawton Scale, which classify the activities performed by the elderly by score. Many of the motor activities present in the scales, such as food, locomotion, personal hygiene, administration of social, economic and self-care functions, were verified through the answers.

The exploration of a tool of easy and fast applicability by health professionals, to evaluate the functionality of the elderly is relevant. However, no similar studies were found, including the same sampling of healthy elderly and correlating with the same variables studied. This study was based on methodological rigor, quality of description and application of the tests, as well as selection and inclusion criteria in the research and collection of information to characterize the sample. However, two possible limitations should be considered. The first refers to the size and type of the sample. The small number of subjects and the fact that the women correspond to almost all of the sample, limits the external validity of the findings. Thus, considering the limitations presented, there are future perspectives for a study of higher sampling, with more homogenous distribution and stratified by sex.

## CONCLUSION

For the sample of healthy elderly and in the proposed experimental conditions, an association was found between HGS, submemal cardiorespiratory capacity and body mobility. Considering the high correlation between these aspects and functionality, it is possible to infer that HGS can be a relevant indicator of functional capacity in this population.

## Data Availability

Dados desenvolvidos na instituicao pelo Grupo de estudos em Reabilitacao (GEReab).

https://gereab.com/

